# Biometric linkage of longitudinally collected electronic case report forms and confirmation of subject identity: An open framework for ODK and related tools

**DOI:** 10.1101/2022.10.13.22280647

**Authors:** Chrissy h Roberts, Callum Stott, Marianne Shawe-Taylor, Zain Chaudhry, Sham Lal, Michael Marks

## Abstract

The availability of low-cost biometric hardware sensors and software makes it possible to rapidly, affordably and securely sample and store a unique and invariant biological signature (or biometric “template”) of participants in research and trials. This has applications in consent, linkage of case reporting forms collected at different times, and in confirmation of participant identity for purposes of safety monitoring and adherence to international data laws.

The use of mobile electronic data collection software has recently become commonplace in clinical trials and research. A raft of tools based on the open-source ODK project now provide diverse options for data management that work consistently in resource-restricted settings, but none have built-in functionality for capturing biometric templates.

In this study, we report the development and testing of a novel open-source app and associated method for capturing and matching biometric fingerprint templates during data collection with the popular data platforms ODK, KoBoToolbox, SurveyCTO, Ona and CommCare.

Using data from more than 1000 fingers, we show that fingerprint templates can be used to link data forms with high accuracy and that this accuracy increases with the addition of multiple fingerprints on each data form. By focussing on publishing open-source code and documentation, and by using an affordable (<£50) and mass-produced model of fingerprint sensor, we are able to make this platform freely available to the large global user community that utilises ODK and related data collection systems.

## BACKGROUND

Access to official identification credentials is a crucial step in providing access to (among others) social and fiscal services, political franchise, free travel, birth registration and social engagement. An estimated one-billion people, and more than half of all people living in low- and middle-income countries (LMICs), possess no form of officially recognised identification credentials (1). Identifying, and later confirming the identity of individuals living as members of what The World Bank described as the ‘invisible billion’ is a substantial challenge not only to excluded individuals and communities, but also to governments and other agencies acting in the interest of excluded communities. Within the sphere of public health work carried out in locales where official identification credentials are not available, there have been historical challenges when agencies have needed to identify and to later confirm the identity of participants/subjects of research, clinical trials and interventions.

Clinical trials provide a powerful use-case for the development of novel systems for identifying and confirming identity of stakeholders from the invisible billion. Trials commonly aim to demonstrate the [a] efficacy and [b] safety of a public health intervention; outcomes that with rare exception require data collection to be longitudinal, heterogeneous, asynchronous and complex. The veracity of trial data is a paramount concern of Good Clinical Practice (GCP), the international standards for conduct and reporting of trials; but no gold standard method exists for confirming participant identities at different stages in the progress of a participant through a trial’s protocol. This raises problems that [a] different case report forms (CRFs) which nominally relate to the same participant might in fact derive from two or more distinct individuals and [b] serious adverse events (SAEs) could be inappropriately attributed (or not) to the trial intervention. The first problem compromises the study design and conclusions, whilst the second could lead to the early halting (or inappropriate continuance) of the trial, false positive/negative results and serious patient, community or public health harms.

Such studies are bound by a range of national and international data laws such as the General Data Protection Regulation (EU, 2016/679). Among other laws of similar effect, GDPR (2) protects the data of participants (also known as “data subjects”) from improper processing, whilst simultaneously enshrining in law their broad-ranging legal rights to access, edit or request the erasure of data about them, including data derived from any information and biological specimens they may have provided during their participation. Thus, researchers must be able to effectively identify any participant and link them to their data in order to comply with requests for data deletion.

Digital authentication mechanisms based on biometric data have been proposed as a potential cornerstone in the future of inclusive and trusted identification methods (1). The recent availability of low-cost biometric hardware sensors and software makes it possible to rapidly, affordably and securely sample and store, alongside clinical trial (or other) data, a unique and invariant biological signature (or biometric “template”) of a study participant. By later being able to compare any two templates (i.e. from different CRFs), records can be linked and the identity can be confirmed in a way that is quantifiable, reproducible and automatable. Fingerprint templates are well described data entities that are regulated in the international standard ISO 19794-2:2005 (3,4) and the interoperable ANSI INCITS 378-2004 standard. Templates do not take the form of a photograph or image of a recognisable fingerprint; rather they are an encoded text representation of the characteristic features of the fingerprint. Although iris (5), ear (6) and other sources of biometric templates have been explored and may be preferable in some contexts, fingerprints remain the most familiar form of biometric datum.

The development of novel biometrics systems has taken place alongside the rapid development of electronic data collection (EDC) systems. Historically, clinical trials relied on the use of paper CRFs, but over the last 15-20 years, there has been a widespread shift towards the use of electronic systems. The rate of adoption of EDCs in research and trials has been globally uneven, partly because of the relatively slower development of web infrastructure and limited penetration of the internet in some countries, regions and communities. A raft of EDC tools aimed specifically at work in LMICs have recently become available. These predominantly app-based tools, which include ODK (7), Ona Data (8), KoBoToolbox (9), SurveyCTO (10), CommCare by Dimagi (11) Enketo (12), REDCap (13), OpenClinica (14) and DHIS2 (15) all have the capability to function without reliable internet connections and can be used to generate digitised data sets of very high quality in even the most inhospitable and complex settings (16). Several of these platforms (ODK, SurveyCTO, KoBoToolbox, Ona and CommCare) are derived from a common code-base (ODK) and have overlapping functions, but none of these software platforms currently provide a robust built-in method for capturing verifiable biometric proof of identity data that can be used to later [1] link CRFs that were collected at different times or [2] confirm the identity of a participant requesting deletion of their data.

Low cost and portable biometrics sensors, used in combination with electronic data collection tools such as ODK, provide a potential solution to the problems of participant identification and CRF linkages. In this study, we report the development of a novel open-source app and associated method for capturing and matching ANSI INCITS 378-2004 fingerprint templates during data collection with ODK and related data tools. By focussing on the use of predominantly open-source code with an affordable (<£50) and mass-produced model of fingerprint sensor, we make this platform freely available to all global users of ODK, Ona, KoBoToolbox, SurveyCTO and CommCare.

## METHODS

### Development of the ODK Biometrics Framework

The novel biometrics system consists of two components. The first component is “Keppel”, a smartphone app designed to run on Google Android operating systems. This app provides an input/output (I/O) interface between the ODK Collect app and an ANSI INCITS 378-2004 compliant electronic fingerprint reader/sensor device. The Keppel smartphone app was designed using Android Studio and Software Development Kit (SDK) (17). The initial version of the app works with the low cost Mantra MFS100 Biometric C-Type Fingerprint Scanner (18), functionality for which was based on code templates provided within the Mantra MFS100 Software Development Kit (19). The Keppel app was designed with a view to making the addition of further biometric sensors relatively simple through the implementation of a common scanner interface in the code-base. A software ‘demo’ scanner, which returns simulated fingerprint data, is also included. This allows users to test their fingerprint supported ODK forms without having a scanner connected.

The second component of the system is the Keppel Command Line Interface (CLI), a Java application designed to run on the command line of a desktop, laptop or server-based workstation. The Java application is able to compare any two ANSI INCITS 378-2004 fingerprint templates and to generate a simple unitless score (S) which describes the overall similarity between the two templates. The Keppel Java CLI utilises an existing open-source library (20) to calculate the similarity between fingerprint templates. Calls to the CLI take the form ‘keppel match -p [template1] [template2]’ where [template 1] and [template 2] are plain text copies of the fingerprint templates of interest. The CLI returns either the absolute value of S (i.e. 193), a logical test of whether S is greater than a provided threshold (T) (i.e. matched/mismatched), or both (i.e. matched_193). The code repository for the project is available on GitHub (21)

### Compatibility

We tested the Keppel app with current releases of data collection apps including ODK Collect (v2022.3 Beta 0), SurveyCTO Collect (v2.72), CommCare (v8) and KoboCollect (v2022.1.2). The Keppel CLI was tested on Ubuntu (Linux) v18.04.4, Darwin (Unix) v21.5.0 and Windows Subsystem for Linux (WSL) 2.

### Data collection

Data were collected on encrypted Android handsets using ODK Collect (16) and were hosted at institutional data centres. We recruited volunteers to provide fingerprints for a real-world evaluation of the system. Participants provided their age, gender, skin-tone (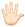 light | 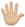 medium-light | 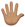 medium | 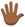 medium-dark | 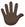 dark), natural level of skin moisture (very dry | dry | normal | moist | very moist), skin greasiness (not greasy | mildly greasy | greasy | very greasy), and overall condition of skin (very bad | bad | good | very good).

Participants were then asked to sequentially scan each of the fingerprints of their right hand on the fingerprint reader. They then repeated this process, recording the same fingerprint a second time to create a matched pair of templates for each of their five fingers. For each of the five fingers, participants indicated whether that finger had visible rough patches (yes/no) and/or smoothed areas (yes/no). This set of matched template pairs represented the true positive group in the analysis. To establish a true negative group, the first five fingerprint templates of each individual were paired with the first five fingerprint templates of another individual from the study. In each case, we ensured that the thumb, index, middle, ring and pinkie prints from the right hand of individual A were paired to the same fingers of individual B’s right hand.

### Analysis

We aimed to recruit a minimum of 200 individuals, generating more than 1000 fingerprints to include in our initial evaluation. Linear regression was used to identify skin characteristics that were associated with changes in the average score among the true positive template pairs. The Receiver Operator Characteristic (ROC) was used to assess the overall performance of the matching algorithm and to identify suitable threshold values for determining matched and unmatched classifications. We used the area under the ROC curve (AUC) as a generalised indicator of the matching algorithm’s performance. The AUC value is an indicator of the aggregate performance of a binary diagnostic assay across all possible classification thresholds. AUC = 1 indicates a perfect classifier and AUC = 0.5 indicates a classifier that performs no better than a random coin-toss. AUC provides a simple way to compare the performance of two diagnostics in a way that is independent of scale and threshold values, and when comparing several classifiers, the one with the higher AUC is generally the more performant assay. In order to determine whether the accuracy of matching could be increased by using more than one finger in the classifier, we calculated a multi-finger score (ΣS) by summing the values of S from two (thumb, index), three (thumb, index, middle), four (thumb, index, middle, ring) or five (thumb, index, middle, ring, pinkie) fingers. ROC analysis was performed and the optimal assay was chosen on the basis of maximising the AUC whilst minimising the number of fingers used to perform the test. All analysis was performed in R v4.2.0.

## RESULTS

All code and working software releases of the app and CLI (v0.3) are provided in Supplementary File 1. The most recent version of the app and CLI are available from our code repository (21).

### Template matching

Overall, 1,010 true positive fingerprint template pairs (n = 2,020 individual templates) were provided by 202 consenting participants with different skin tones, qualities and conditions (Table 1). 1,010 true negative pairs were synthesised as described above. The mean score for a comparison between templates of a true positive pair was S = 163.0 (median 159.4, min 0.6, max 472.7, IQR 97.0 - 219.5). For true negative template pairs, the mean score was S = 5 (median 3.6, min 0.0, max 31.3). ROC analysis of 1,010 true positive and 1,010 true negative template pairs revealed an area under the curve (AUC) of 0.99 (Figure 1A). At the threshold score (T) of T = 27, the fingerprint matching algorithm had a false positive rate of 0.001 and true positive rate of 0.95. A threshold of T = 27 was subsequently used as the cut-point between mismatched (S < 27) and matched (S >= 27) template pairs. A total of 52 (5.14%) true positive pairs were falsely classified as negative. Among the true negative template pairs, just one pair returned a false positive result (S = 31.26).

**Table 1:** Characteristics of study participants and their skin.

| Group | Subgroup | n | % |
| --- | --- | --- | --- |
| Age Group | 20-30 | 104 | 51.5 |
|  | 31-50 | 85 | 42.1 |
|  | 51+ | 13 | 6.4 |
| Skin Tone | dark | 3 | 1.5 |
|  | medium dark | 8 | 4.0 |
|  | medium | 32 | 15.8 |
|  | medium light | 59 | 29.2 |
|  | light | 100 | 49.5 |
| Skin Moisture | very dry | 6 | 3.0 |
|  | dry | 72 | 35.6 |
|  | normal | 110 | 54.5 |
|  | moist | 14 | 6.9 |
| Skin greasiness | not greasy | 105 | 52.0 |
|  | mildly greasy | 85 | 42.1 |
|  | greasy | 12 | 5.9 |
| Skin Condition | bad | 21 | 10.4 |
|  | good | 157 | 77.7 |
|  | very good | 24 | 11.9 |
| Creams or lotions used today | no | 109 | 54.0 |
|  | yes | 93 | 46.0 |
| Oils or emollients used today | no | 183 | 90.6 |
|  | yes | 19 | 9.4 |
| Medication used today | no | 195 | 96.5 |
|  | yes | 7 | 3.5 |
| Hand sanitiser used today | no | 48 | 23.8 |
|  | yes | 154 | 76.2 |
| Surgical scrub used today | no | 106 | 52.5 |
|  | yes | 96 | 47.5 |

**Figure 1:**
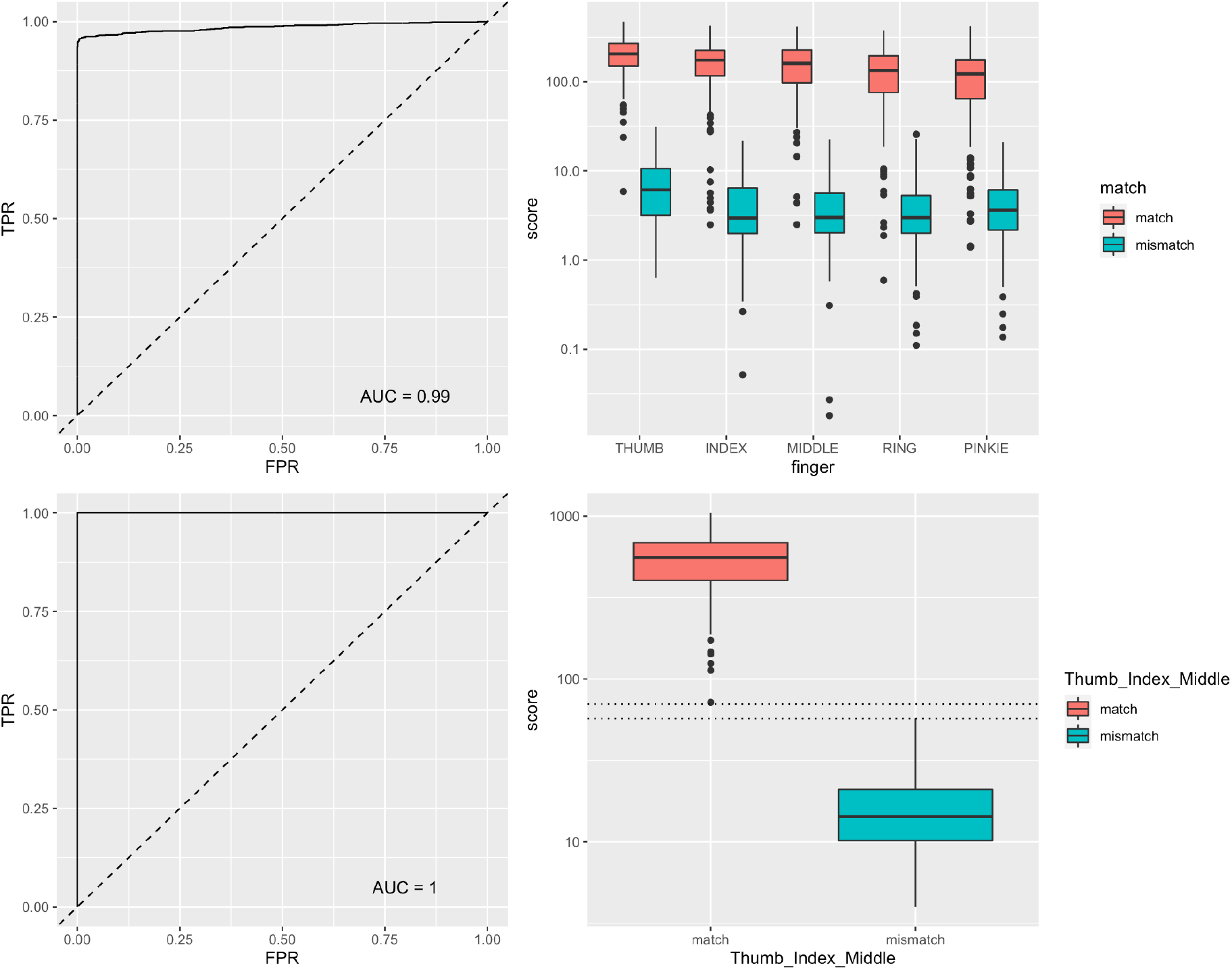
**A**. ROC analysis (Single Finger). The area under the ROC curve for any pair of fingerprint templates was 0.99. This analysis was based on 1,010 true positive and 1,010 true negative template pairs. **B**. Assay performance variations by finger. There was a progressive (from thumb to pinkie) decrease in the average score of a true positive template pair according to which finger was used. **C**. ROC analysis (score summed across thumb, index and middle finger) The area under the ROC curve was 1.00, indicating a perfect delineation between true positives and true negatives. **D**. Summed scores of three-finger scanning among true positive and true negative groups.

The majority of participants (n = 170, 84.16%) received positive matches (S > 27) for all five fingers. Of the remaining 32 participants, 18 (8.91%) received positive matches for four fingers, with nine (4.46%), two (0.4%) and one (0.1%) participants receiving positive matches at three, two or one fingers respectively. Compared to the average values for thumb templates, relative values of S (RS) were significantly and progressively lower for template pairs representing the index (RS = -34.2, SE = 8.0), middle (RS = -43.7, SE = 8.1), ring (RS = -68.6, SE = 8.1) and pinkie (RS = -87.0, SE = 8.1) fingers (Figure 1B, Table 2).

**Table 2:** Factors associated with increased or decreased scores.

| Factor | RS* | SE** | P |
| --- | --- | --- | --- |
| (Intercept) | 228.19 | 23.96 | <0.001 |
| Thumb (Ref) | REF | REF | REF |
| Index Finger | -34.24 | 8.04 | <0.001 |
| Middle Finger | -43.71 | 8.06 | <0.001 |
| Ring Finger | -68.64 | 8.05 | <0.001 |
| Pinkie Finger | -86.95 | 8.06 | <0.001 |
| Rough Areas | -16.89 | 12.06 | 0.162 |
| Smoothed Areas | 1.20 | 7.17 | 0.867 |
| Age 20-30 | REF | REF | REF |
| Age 31-50 | 9.28 | 5.56 | 0.095 |
| Age 51+ | -70.86 | 11.85 | <0.001 |
| Skin Tone: Dark | REF | REF | REF |
| Skin Tone: Medium Dark | -11.46 | 26.45 | 0.665 |
| Skin Tone: Medium | -23.40 | 23.53 | 0.320 |
| Skin Tone: Medium Light | -16.52 | 22.88 | 0.470 |
| Skin Tone: Light | -14.62 | 22.69 | 0.520 |
| Skin Moisture: Normal | REF | REF | REF- |
| Skin Moisture: Moist | 1.03 | 11.02 | 0.925 |
| Skin Moisture: Dry | -0.79 | 5.90 | 0.893 |
| Skin Moisture: Very Dry | 17.24 | 16.37 | 0.292 |
| Skin: Not Greasy | REF | REF | REF |
| Skin: Mildly Greasy | 5.97 | 5.65 | 0.290 |
| Skin: Greasy | -0.39 | 11.74 | 0.974 |
| Skin Condition: Good | REF | REF | REF |
| Skin Condition: Bad | -6.63 | 9.00 | 0.462 |
| Skin Condition: Very Good | 26.43 | 8.62 | 0.002 |
| Applied Lotion/Cream Today | -11.59 | 5.86 | 0.048 |
| Applied Emollient/Oils Today | -10.21 | 10.04 | 0.309 |
| Applied Medication Today | 3.05 | 16.80 | 0.856 |
| * RS : The average relative value of the matching score S, compared to the reference group (REF) for factors with multiple responses, or compared to the counterfactual for factors with binary responses. |  |  |  |
| ** SE : Standard Error |  |  |  |

Average relative scores for true positive template pairs were lower when collected from people aged over 50 (RS = -70.9, SE = 11.9) and higher when collected from people who described their overall skin condition as ‘very good’ (RS = 26.4, SE = 8.6). There appeared to be no variation in the average score of a true positive template pair on the basis of skin tone, moisture level, or greasiness (Table 2). There was some evidence for a slight reduction in scores when the participant had applied creams or lotions to their hands on the day of sampling (RS = -11.6, SE = 5.9) although this did not appear to be confirmed among those who had applied oils, emollients or medications. The presence of rough or smooth patches on the fingertips did not appear to affect the average score (Table 2).

There was a 94.9% probability that any two fingerprint templates taken from a single finger would return a positive match (defined as S > 27). The probability that there would be at least one positive result among any two template pairs taken from two fingers of the same individual was 99.0%. Using three, four or five fingers, the probability of returning at least one positive result (S > 27) rose to 99.6%, 99.8% and 100% respectively.

Using the sum of the scores from two (AUC = 0.997), three (AUC = 1.0), four (AUC = 1.0) or five (AUC =1.0) fingers indicated a general improvement in the performance of the matching algorithm with an increasing number of fingers used. In this data set, we found that a perfect classifier (AUC = 1) could be achieved when three fingers (thumb + index + middle) were used (Figure 1C, 1D). In the three-finger model, the mean summed score for a true positive was ΣS = 557.18 (median 557.18, min 71.94, max 1046.43, IQR 402.79-686.03). For true negatives, the mean summed score was ΣS = 16.53 (median 14.3, min 3.99, max 57.22) (Figure 1D). The gaps between the highest values of a true negative and the lowest values of a true positive in each classifier were 13 (three fingers), 12.3 (four fingers) and 62.7 (five fingers) units. This indicated that among the three classifiers where AUC=1.0 (i.e. the models where three, four or five fingers were combined), there was an additional performance gain from using more fingers, as the separation between the positive and negative groups (and consequently the proportion of all possible threshold values where the classifier worked perfectly) increased.

### System Compatibility and performance

The CLI was highly performant. Using a single CPU on a 2019 MacBook Pro (2.3 GHz, 8 Core Intel i9, 64 GB RAM), we compared 400 pairs of templates in 140 s, which equated to 0.350s per comparison. Using a 16-core parallel implementation through the R package “furrr” (22), we processed 2000 template pairs in 69.7 s, equating to 0.035 s wall-clock time per comparison. The Keppel CLI worked as expected on Linux, Unix and within the Windows Subsystem for Linux. We tested compatibility of the Keppel app with several commonly used data collection apps that are based on ODK’s code. We found that our biometrics app functioned as expected with ODK Collect (v2022.3 Beta 0), SurveyCTO Collect (v2.72) and KoboCollect (v2022.1.2). Although we anticipate that the system can work with CommCare, we were not able to confirm compatibility with a current version of the CommCare app (v8) because required functions (external app integration) were paywalled in CommCare, only being made available for higher tier subscribers of that service. Ona does not have a standalone data collection app and users instead connect to Ona servers via ODK Collect, therefore making it compatible with the Keppel app. The system is currently not compatible with Go.Data, DHIS2 or OpenClinica.

## DISCUSSION

Scant literature addresses topics of participant (mis)identification in research and trials, but in clinical care the subject is better explored (23,24) and highlights that patient mis-identification is common even in high income countries (HICs) with well-developed health information systems. In resource-restricted settings, challenges are magnified, particularly when complex longitudinal data collection centres around the use of experimental medical products, or the surveillance of infectious diseases in remote locales (16).

The current generation of mobile data collection software is well developed for use in these settings, with off-grid functionality the norm; but no system historically provided a simple to use method to support participant identification. We have created open-source code and related applications that introduce options for fingerprint template capture, and workstation-based template matching when collecting data with ODK’s native smartphone app ODK Collect. We have tested and confirmed that the app also works with the majority of other software that have been forked (copied and further developed as separate software, usually by an independent developer group) from the ODK project. This includes KoBoToolbox, Ona and SurveyCTO, which together with CommCare (compatibility unconfirmed) and ODK have a dominant market share of mobile data collection tools used in public health, humanitarian activities, election monitoring and elsewhere. The system is not currently compatible with major data collection systems that are not based on ODK’s code (i.e. OpenClinica, DHIS2, REDCap and Go.Data).

There appeared to be several factors that significantly influenced the quality of template matching. Most importantly, we highlighted that for the purposes of clinical trials, it was insufficient to rely on scanning just one fingerprint template in each CRF. The data presented here indicate that the matching of a single fingerprint template pair has absolute accuracy of around 95%. This is perhaps acceptable for confirming identity during the process of a data subject access request, where any issues of false-negative results might be rectified in most cases by trying again with a new scan from the same finger. 95% accuracy is meanwhile too low for the practical purposes of needing to confirm links between data. A recent clinical study protocol aimed to vaccinate 500,000 participants (25). Assuming 10 CRFs per participant and single scanning of fingerprint templates on each CRF, then we estimate that there would be 5,000,000 comparisons, of which around 250,000 linked records would not match because of the 5% error rate. At such scales, subtle (i.e. 3-4 decimal places) increases in classifier performance could translate into substantial decreases in the actual number of errors in a real world setting. By using multi-finger scanning (and then summing the matching scores), we reduced the error to levels that were undetectable in our test data. We also saw that whilst our relatively small data set was underpowered to discern the subtle performance increases in AUC of a ROC analysis when four or five fingers were used (compared to three fingers where AUC = 1), there was evidence from the growing separation of the negative and positive populations that using more fingers would continue to decrease error in the system and improve accuracy. We therefore recommend that all users of the system should collect a minimum of three and ideally more fingerprint templates in each CRF. We found that the average score for a matched pair was highest for thumb templates and that each subsequent finger moving towards the pinkie had a lower average score. When choosing which fingers to scan, it may be valuable to prioritise fingers accordingly.

Certain participant characteristics influenced the probability of obtaining a high matching score from a pair of templates collected from the same finger. We found that template pairs from those who self-described as having ‘very good’ skin condition were more likely to produce high scores. The score neither appeared to be influenced by skin tone or greasiness, nor by the presence of rough or smooth areas. Template pairs from people aged over 50 were substantially lower (RS = -70) than those from people aged 20-30. In this study, we focused on the use of fingerprint templates taken from the right hand and we are therefore unable to determine whether there could be performance differences when using templates from the left hand.

The Keppel app and CLI is not the first biometrics platform that is compatible with the ODK ecosystem. Simprints (26) is well-developed software which is compatible with ODK-like systems, as well as with DHIS2. This platform openly shares code (27) but its use may require a managed account provided by the not-for-profit Simprints Technology Ltd. Use of Simprints also appears to require access to proprietary scanner hardware, access to which may also require direct collaboration with the company. Whilst Simprints does have wider functionality than the system presented here (including contactless biometrics and on-device matching/verification), our project is novel in being designed around low cost, off-the-shelf hardware and a fully “open software - open documentation” model which makes setup simple, which can be used by any project using compatible software and which does not require support from, partnership with, or subscriptions provided by any third party.

Most trials have tens, hundreds or thousands of participants, meaning that compute times for the Keppel CLI are negligible in most cases, even when multi-finger scanning and multiple CRFs are used. The CLI was however highly performant at scale and we predict that in a very large trial with 15,000,000 templates (500,000 participants, 10 CRFs, three-finger scanning) the CLI could process all required template comparisons in around one week of wall-clock time on a higher-end consumer laptop (2.3 GHz, 8 Core Intel i9, 64 GB RAM). Using a high-performance cluster would reduce this in proportion to the number of CPUs available (for instance, wall clock time would be around 15 minutes if using a 96 CPU cluster).

Limitations of this study include that the first release of the Keppel app does not perform on-device matching or verification, and that the app works with just one specific and proprietary hardware scanner device. To ameliorate the risk that the compatible hardware may become obsolete, we encoded the app in such a way that further devices (including sensors for other biometrics) could be added with relative ease. Our open-source code is also available for community-led maintenance and further development. This study was performed during the COVID-19 pandemic at a time when collecting fingerprints was a non-trivial process. As such, we did not achieve a sample size that would have allowed us to estimate with greater precision the level of error that remained when performing four- or five-finger scanning. Our error estimates are therefore indicative and should be interpreted only to show that multi-finger scanning is the preferable method of sampling. We were also unable to include children and infants in the study (because of risk assessments related to COVID-19) and are consequently not able to use these data to assess whether the system performs well in young and very young people. This will be a topic of future study.

The use-case for these tools in clinical trials, medical informatics, research and health care is extensible to the broader use-cases of humanitarian electronic data management tools. Central to the value of low cost and reliable bioinformatics tools in action for public good are deployments that will address the problems of the excluded “missing billion”. We anticipate that the tools presented here, along with those which will emerge from continuous open-source developments of the code-base, will contribute to efforts that will provide official and semi-official identification credentials; thereby enhancing political franchise, supporting free and fair elections, enabling birth registration & census and providing solutions to many other personal identification challenges in resource-restricted settings.

## Data Availability

The latest release of the app and CLI, along with all code relating to this work, are available at the project home page: https://github.com/LSHTM-ORK/ODK_Biometrics. Copies of all code and release version 0.3 are provided as supplementary data. The Keppel App runs on Android Devices. The app works in combination with one of ODK Collect, KoBo Collect and SurveyCTO Collect but may work with other similar apps. The Keppel CLI was programmed in Kotlin and is platform independent. All code is released on the MIT Licence (https://opensource.org/licenses/MIT). At the time of writing, the Mantra MFS100 Biometric C-Type Fingerprint Scanner was widely available from online retailers as well as from the manufacturer www.mantratec.com. Fingerprint data cannot be shared for ethical reasons.

## ETHICS STATEMENT

Ethical permission for the study was granted by the London School of Hygiene & Tropical Medicine Observational Research Ethics Committee (Ref. 22562). Before taking part, participants were provided with written information about the study and were able to ask project staff questions about the study before providing verbal and electronic informed consent to take part. The study was fully anonymous and the study team did not keep any record of the name or other personal information about the identities of the participants. In order to allow participants to change their minds and to leave the study at any time, the study information sheets carried unique codes that were stored on the project database. Any participant who wished to leave the study was instructed to contact the project team at any time and to provide the code number(s) of records which would then be permanently deleted.

## DATA AVAILABILITY STATEMENT

The latest release of the app and CLI, along with all code relating to this work, are available at the project home page: https://github.com/LSHTM-ORK/ODK_Biometrics. Copies of all code and release version 0.3 are provided as supplementary data. The Keppel App runs on Android Devices. The app works in combination with one of <u>ODK Collect</u>, <u>KoBo Collect</u> and <u>SurveyCTO Collect</u> but may work with other similar apps. The Keppel CLI was programmed in Kotlin and is platform independent. All code is released on the MIT Licence (https://opensource.org/licenses/MIT). At the time of writing, the Mantra MFS100 Biometric C-Type Fingerprint Scanner was widely available from online retailers as well as from the manufacturer www.mantratec.com. Some code used in this project was based on the Mantra MFS100 Software Development Kit, which at time of writing was available from Mantra Softech https://download.mantratecapp.com/. Fingerprint data cannot be shared for ethical reasons.

## AUTHORS’ CONTRIBUTIONS

ChR, CS, SL & MM Conceptualised the study. CS & ChR wrote and tested the code and documentation. ZC, MST, MM & SL collected the Data. ChR performed the statistical analysis. All authors contributed to writing, reviewing and approving the content of the manuscript.

## ACKNOWLEDGEMENTS

We are grateful to the volunteers who took part in the study, to the technical services team at Mantratec for their support in the use of their SDK and to Eleanor Martins for project administrative leadership. We would also like to acknowledge the value of many conversations about biometrics that we had with members of the ODK Forum (https://forum.getodk.org/) and partners from the Tujiokowe trial consortium.

## REFERENCES

1. Inclusive and Trusted Digital ID Can Unlock Opportunities for the World’s Most Vulnerable. World Bank https://www.worldbank.org/en/news/immersive-story/2019/08/14/inclusive-and-trusted-digital-id-can-unlock-opportunities-for-the-worlds-most-vulnerable [Accessed August 24, 2022]

2. General Data Protection Regulation (EU, 2016/679). https://eur-lex.europa.eu/eli/reg/2016/679/oj

3. ISO/IEC 19794-2 Information technology — Biometric data interchange formats — Part 2: Finger minutiae data. (2011) Second edition: https://webstore.iec.ch/preview/info_isoiec19794-2%7Bed2.0%7Den.pdf

4. Važan R. ISO/IEC 19794-2:2011/Cor.1:2012 Summary. https://templates.machinezoo.com/iso-19794-2-2011 [Accessed May 4, 2022]

5. Corby PM, Schleyer T, Spallek H, Hart TC, Weyant RJ, Corby AL, Bretz WA. Using biometrics for participant identification in a research study: a case report. J Am Med Inform Assoc (2006) 13:233–235. doi: 10.1197/jamia.M1793

6. Ragan EJ, Johnson C, Milton JN, Gill CJ. Ear biometrics for patient identification in global health: a cross-sectional study to test the feasibility of a simplified algorithm. BMC Res Notes (2016) 9:484. doi: 10.1186/s13104-016-2287-9

7. ODK. https://getodk.org/ [Accessed June 23, 2022]

8. Ona Data. https://ona.io/ [Accessed June 23, 2022]

9. KoBoToolbox. https://www.kobotoolbox.org/ [Accessed June 23, 2022]

10. SurveyCTO. (2018) https://www.surveycto.com/ [Accessed June 23, 2022]

11. CommCare by Dimagi. https://www.dimagi.com/commcare/ [Accessed June 23, 2022]

12. Enketo. https://enketo.org/ [Accessed June 23, 2022]

13. REDCap. https://www.project-redcap.org/ [Accessed June 23, 2022]

14. OpenClinica. (2022) https://www.openclinica.com/ [Accessed June 23, 2022]

15. DHIS2. (2018) https://dhis2.org/ [Accessed June 23, 2022]

16. Marks M, Lal S, Brindle H, Gsell P-S, MacGregor M, Stott C, van de Rijdt M, Almazor GG, Golia S, Watson C, et al. Electronic Data Management for Vaccine Trials in Low Resource Settings: Upgrades, Scalability, and Impact of ODK. Front Public Health (2021) 9:665584. doi: 10.3389/fpubh.2021.665584

17. Android Studio and SDK tools. https://developer.android.com/studio [Accessed June 23, 2022]

18. Mantra - Redefining Biometric Security Solution. Mantra Softech India Pvt Ltd https://www.mantratec.com/ [Accessed June 23, 2022]

19. Mantra MFS100 Software Development Kit. https://download.mantratecapp.com/ [Accessed June 23, 2022]

20. Važan R. sourceafis-java: Fingerprint recognition engine for Java that takes a pair of human fingerprint images and returns their similarity score. Github. https://github.com/robertvazan/sourceafis-java [Accessed September 21, 2022]

21. Roberts C, Stott C. ODK_Biometrics. Github. https://github.com/LSHTM-ORK/ODK_Biometrics [Accessed September 21, 2022]

22. Vaughan D, Dancho M. furrr: Apply Mapping Functions in Parallel using Futures. (2022) https://CRAN.R-project.org/package=furrr

23. Riplinger L, Piera-Jiménez J, Dooling JP. Patient Identification Techniques - Approaches, Implications, and Findings. Yearb Med Inform (2020) 29:81–86. doi: 10.1055/s-0040-1701984

24. Solomon RP, Goldberg-Alberts A, Marella WM, Pusey C, Rebold C, Anderson PA, Addis LM, Barndt JL, Drozd E, Fenton A, et al. reECRI Institute PSO Deep Dive - Patient Identification: Executive Summary. https://www.ecri.org/Resources/Whitepapers_and_reports/PSO%20Deep%20Dives/Deep%20Dive_PT_ID_2016_exec%20summary.pdf

25. Watson-Jones D, Kavunga-Membo H, Grais RF, Ahuka S, Roberts N, Edmunds WJ, Choi EM, Roberts CH, Edwards T, Camacho A, et al. Protocol for a phase 3 trial to evaluate the effectiveness and safety of a heterologous, two-dose vaccine for Ebola virus disease in the Democratic Republic of the Congo. BMJ Open (2022) 12:e055596. doi: 10.1136/bmjopen-2021-055596

26. Simprints Technology. https://www.simprints.com/ [Accessed June 23, 2022]

27. Simprints. Github. https://github.com/Simprints [Accessed June 23, 2022]

